# Developing contents for a digital drug adherence tool with reminder cues and personalized feedback: a formative mixed-methods study among children and adolescents living with HIV in Tanzania

**DOI:** 10.1101/2023.03.22.23287383

**Authors:** Iraseni Swai, Lisa Lynn ten Bergen, Alan Mtenga, Rehema Maro, Kennedy Ngowi, Benson Mtesha, Naomi Lekashingo, Takondwa Msosa, Tobias F. Rinke de Wit, Rob Aarnoutse, Marion Sumari-de Boer

## Abstract

Children and adolescents living with HIV (CALHIV) form a significant proportion of people living with HIV (PLHIV). Optimal adherence (>95%) to medication is needed to achieve viral suppression. However, optimal adherence remains a challenge among CALHIV. Digital adherence tools (DAT) like the Wisepill® device have proven feasible among adult PLHIV. Still, there are concerns about unwanted disclosure of HIV status due to content in short message service (SMS) that serve as reminders. We assessed the needs, contents, and acceptability of a DAT intervention among CALHIV.

We conducted a mixed-methods study among CALHIV with their parents/guardians. We performed a survey and then purposively selected participants who used the intervention for one month. They received SMS text reminders that differed over time from asking if the medication was taken to more neutral SMS like “take care”. After one month, participants received tailored feedback based on automatically generated adherence reports. Afterwards, we completed exit interviews, in-depth interviews, and focus-group discussions. We analysed quantitative findings descriptively and used thematic content analysis for qualitative data.

We included 284 participants in the survey and 40 used the intervention. Among participants who often forgot medication intakes, 93% of adolescents and 83% of children’s parents/guardians were interested in receiving reminders. Among participants who used DAT, 90% had good experience receiving reminders and agreed that SMS made them take medication. However, 25% experienced network problems. Participants were happy to use the device. Further, they preferred neutral reminder SMSs that did not mention the word ‘medication’, but preserved confidentiality. Adherence reports inspired good adherence. None of the participants experienced unwanted disclosure or stigmatisation due to DAT. However, 5% of adolescents were concerned about being monitored daily.

This study provided insights on how to customise DAT. We will implement this in a clinical trial to assess effectiveness in improving adherence.

**Author’s summary:** CALHIV are required to take antiretroviral medication on time, every day, for the rest of their lives. That is necessary to suppress the virus and live a healthy life. Maintaining that consistency is not easy. Digital tools that assist in reminding medication time, like the Wisepill device, have proven feasible among adult PLHIV. However, there are concerns about HIV status disclosure due to the contents used in the SMSs. We tested a DAT intervention in which participants used the Wisepill device, received reminder SMS in their phones and adherence reports on how they took medication over a month. We sent SMS contents that differed over time from asking if the medication was taken to more neutral SMS like “take care”. After one month, we asked participants their opinions about the interventions. Most participants were happy to use the device and to receive neutral SMS contents that did not mention ‘medication’ and which preserved their confidentiality. Adherence feedbacks motivated good adherence behaviour. However, some participants experienced network challenges, and 5% of adolescents were concerned about being monitored daily. We will use preferred SMS contents in the clinical trial that will assess the effectiveness of the DAT in improving adherence among CALHIV.

## Introduction

Children and adolescents living with HIV (CALHIV) form a significant part of people living with HIV (PLHIV). Globally, it was estimated that 1.7 million children aged 0-14 years were living with the human immunodeficiency virus (HIV) in 2021, and 88% resided in Sub-Saharan Africa (SSA).(1,2) Tanzania was mentioned among the top six countries in SSA with the highest number of CALHIV aged 0-19 years, with 96,000 children aged 0-14 years living with HIV and only 60% of them on antiretroviral therapy (ART).(1,3)

The UNAIDS’ 95% fast-track treatment goals aim to end the HIV epidemic by 2030 by having 95% of PLHIV know their status, 95% of those who know should have initiated ART, and 95% of those on ART to have suppressed viral loads.(4) The introduction of combination ART has significantly changed the face of HIV/AIDS by reducing morbidity and mortality rates. Good adherence to ART is needed to achieve the last 95% goal of sufficient viral suppression.(5–8) Adherence to treatment poses a significant challenge for people living with HIV since treatment is for life.

Several studies have shown that CALHIV are less likely to have a suppressed viral load (≤ 200 copies/ml) compared to adults due to poor medication adherence (< 95%) and poor retention in care.(9–12) A recent study done in Mwanza, Tanzania, reported that only 62.5% and 65.3% of children and adolescents, respectively, reached optimal adherence (> 95%) by pharmacy refill (13). In contrast, in another study in Dar es Salaam, 37% of adolescents could not reach optimal adherence.(14) Because HIV infection requires lifelong treatment, CALHIV face more years of needed optimal adherence than adults. Moreover, children depend primarily on their parents or caregivers regarding disease management.

Multiple barriers that hinder adherence have been reported by CALHIV. Children and adolescents in boarding schools face particular adherence barriers like fear of status disclosure mainly due to drug packaging, lack of privacy during medication intake time, challenges with drug storage and lack of a structured support system in schools.(15–17) Among adolescents, the mentioned barriers were stigma, adverse effects of ART, lack of assistance, depression and forgetfulness.(15,16,18,19) Reported factors among caregivers for CALHIV are forgetfulness, long clinic waiting hours, busy schedules and limited knowledge about HIV.(20, 21) Therefore, developing interventions to overcome adherence barriers is of utmost importance to maintain optimal viral suppression.

In recent years, interest has grown in using digital adherence tools (DAT) with reminder cues through short message services (SMS) for improving adherence to treatment and retention in care.(22–27) Several studies have found DATs with reminder SMS feasible among adult PLHIV.(28–31) However, there is limited evidence of its effectiveness among CALHIV in improving adherence.(32–34) Also, there are concerns about unwanted disclosure of the HIV status due to the wording used in the SMS texts.(35–40)

Tanzania has high penetration of mobile phones (89%), we believe that phone-based interventions can potentially assist CALHIV with taking their medication.(41) Through DAT interventions, healthcare workers can monitor medication adherence with access provided through online adherence reports and subsequently provide personalized adherence feedback to the individual. Hence, whereas the DAT device only addresses the barrier of ‘forgetfulness’ by sending SMS reminders, personalized feedback using adherence reports during clinic visits can address specific barriers faced by CALHIV.

This study aimed to (1) assess the needs and contents for an SMS based DAT and (2) assess the acceptability of the DAT intervention (i.e. using the device, receiving different SMS contents and personalized adherence reports). Therefore, we pilot-tested a DAT intervention with different SMS contents and personalized feedback that aims to tackle a wide range of concerns reported by CALHIV. The results of this formative study will inform the customization of the DAT intervention that we will test in a future randomized controlled trial (RCT).

## Methods

### Study design

In this formative study, we used a convergent parallel mixed-methods design. We collected quantitative and qualitative data concurrently and analysed them independently to answer our study objectives.(42) The study consisted of a survey and a one-moth DAT intervention in a selection of participants, followed by a semi-structured exit interview, in-depth interview (IDIs) and focus-group discussions (FGDs) among those who had received the DAT intervention. The study was approved by the College Research and Ethical Review Committee (CRERC) of Kilimanjaro Christian Medical University College (KCMUCo) and the National Health Research Ethics Sub-Committee (NatHREC) of the National Medical Research Institute (NIMR) of Tanzania.

### Study population and study area

We conducted the study in Kilimanjaro Region, Tanzania. Kilimanjaro Region is in the north-eastern part of Tanzania and borders Kenya. We recruited eligible patients from five health facilities: a referral hospital Kilimanjaro Christian Medical Centre (KCMC), the regional Mawenzi Hospital, the district hospital Kibosho Hospital and Majengo and Pasua Health Centre. Children (aged 0 – 14 years) with their caretakers and adolescents (15 – 19 years) living with HIV were eligible if they met the following inclusion criteria: (1) attending Care and Treatment Centres (CTC) in one of the five health facilities, (2) willing to use the DAT and receive SMS, (3) having received ART for at least six months, and (4) having a mobile phone with a registered SIM card (for children this was the caregiver). Exclusion criteria were admission to a hospital at study entry or previous or concurrent participation in other digital adherence trials. We provided phones for those lacking one.

### Sample size

There was no information on how many participants experienced barriers to using DATs. Therefore, to estimate the sample size, we used the arbitrary percentage of 50%, with **α**=0.05 and a power of 80% (estimation of single proportion sample calculation). This resulted in a sample size calculation of 142 children and their caregivers and 142 adolescents participating in the survey. Among those, we purposively selected 20 children and 20 adolescents to participate in the DAT intervention. We considered twenty participants adequate to reach data saturation.

### Study procedures

We used an extensive procedure with guardians/parents to consider whether the HIV status was disclosed to the child and asked for assent from children older than eight years who were aware of their status. For participants < 18 years, we obtained written informed consent from their caregivers or parents. We obtained written informed consent from adolescents aged 18 to 19 years. We conducted a second written informed consent procedure for participants enrolled in the DAT intervention.

### Survey

We used convenience sampling to approach 142 participants for the survey. We collected data from September 2021 until March 2022. Using a semi-structured questionnaire, we recorded demographic characteristics, ART regimen, time of usual intake, adherence levels, the need for SMS reminders to take medication, frequency and timing of the SMS and barriers to using SMS. We also recorded information about owning and using mobile phones, experience with communicating through SMS, network issues and other phone-communication challenges.

### The Digital adherence tool (DAT) intervention

We applied purposive sampling when selecting 20 participants for the DAT intervention. We purposively selected children with their parents/guardians and adolescents based on different ages, varied adherence levels and different health facilities. Participants used our DAT for one month to understand the device’s mechanism and receive different reminder messages.

After one month, participants received tailored feedback based on their adherence reports automatically generated by the DAT. Nurse Counsellors performed the feedback session at the preferred HIV clinic of the participant. The nurse counsellor discussed adherence levels and challenges to adherence with the participant to eventually agree on an action plan with set adherence goals for the next clinic visit. The trans-theoretical model (TTM) of behavioural change, also called the Stages of Change Model, was applied to shape the intervention as practical guidance during the feedback consultation.(43) Consequently, during one feedback session, respondents moved through six stages of change, from awareness of poor adherence and intention to change behaviour towards establishing an action plan to improve adherence.

We collected data in Swahili by trained research assistants not part of the participants’ standard HIV care. Following the feedback consultation, participants attended a short exit interview. Then, in-depth interviews (IDIs) and focus-group discussions (FGDs) were conducted depending on participant availability.

We used the Wisepill® RT2000 pillbox for the DAT intervention, an internet-enabled medication dispenser. The device registers each time it is opened and automatically generates a report indicating the time of intake per day. It contains a rechargeable battery that can run longer than six months if fully charged. The device uses a subscriber identity module (SIM) card and a GSM communication chip. When the dispenser is opened, it sends an electronic medication record to a central management system (Wisepill® Web Server) through the GPRS data network. If patients forget to take their medication at the prescribed time, the system sends a text message to their mobile phones. We sent reminder SMSs, firstly half an hour before intake time and secondly, one hour after the usual time of intake if the participant forgot to open the pillbox at the prescribed intake time. In the first week, we used general SMS content, like: *“Don’t forget to take your medication!”* We adopted this content from our previous study.(44) In the second, third, and fourth week more neutral content was used, such as *“Remember your health”, “The time is near”,* or *“Remember!”*

Healthcare providers and the research team monitored the patients’ medication events through online authorized access displaying a graph report regarding the medication intake of patients. The nurse discussed this report with participants during adherence feedback sessions at the end of the intervention.

### Exit-interview

We used a semi-structured questionnaire for the exit interview at the end of the DAT study. We designed the questionnaire to collect information on the participant’s adherence to ART medications, i.e. the date and number of dispensed, swallowed and missed pills. We asked participants’ opinions about the appearance and functionality of the device, their preference of the received SMS contents, to which participants responded with either ‘Yes’, ‘No’ or ‘I do not remember‘, and their perspective on the adherence feedback with the nurse councillors.

### In-depth interviews (IDIs)

We applied a semi-structured topic guide for the IDIs with those participating in the DAT study. We interviewed participants to obtain more in-depth information about issues regarding the use of the Wisepill® device, the SMS frequency, SMS content and the feedback consultation. Moreover, we collected information on the potential of using educational or motivational SMS. We measured acceptability using the Theoretical Framework of Acceptability.(45) We adapted the topic guides during the research process in an iterative manner.

### Focus-group discussions (FGD)

We conducted FGDs amongst participants in the DAT study to verify information obtained from IDI by gaining insights into community views. We conducted FGDs among different groups of children, adolescents, and caregivers. Each group involved 6-8 participants that we purposely selected based on their different opinions of the intervention. We used the interview guide in an interactive process. We transcribed interviews verbatim and then translated them into English.

### Theoretical Framework and Data analysis

For the acceptability of the DAT intervention, we applied the Theoretical Framework of Acceptability (TFA) post-intervention.(45) The TFA is defined as a multi-faceted framework that reflects the extent to which people delivering or receiving a health care intervention consider it appropriate, based on anticipated or experiential cognitive and emotional responses to the intervention. We applied all seven TFA constructs, which are (1) *Affective attitude*, whether CALHIV were satisfied with the DAT intervention; (2) *Intervention coherence*, the extent to which CALHIV understood the DAT and how it worked; (3) *Perceived effectiveness*, the extent to which the DAT intervention increased CALHIV’s motivation to be adherent to ART; (4) *Burden,* the amount of effort that was required to participate in the DAT intervention; (5) *Ethicality*, the extent to which the DAT had a good fit with an individual’s value system, such as religion, beliefs, rights and culture; (6) *Opportunity costs*, the benefits, profits or values that were given up to engage in the DAT intervention; (7) *Self-efficacy*, the CALHIV’s confidence that he or she can perform the behaviour required to participate in the DAT.

We entered quantitative data from the survey and exit interviews in REDCap® (Research Electronic Data Capture), an open-source secure web application for building and managing online surveys and databases.(46)

To answer the two study objectives, we computed descriptive frequencies of variables to give an overview of the data from the survey and exit interviews. We analysed data using SPSS® Statistics 27. For data that were not normally distributed, we reported outcomes as medians and the interquartile range (IQR) to indicate the measure of dispersion.

Furthermore, for the IDIs and the FGDs, we conducted a thematic framework analysis using the TFA to gain more insight into the context around the needs and contents for reminder SMS and the acceptability of using the DAT intervention. We applied inductive and deductive coding. We first summarized transcripts to get familiar with the data. IS, LB and AM created memos based on the first six interviews and we developed a preliminary codebook. We organized data inductively into each TFA construct. We discussed the codebook, and we then applied codes to the remaining transcripts. We used NVivo version 12 pro for data organization.

## Results

### Number and characteristics of participants in the sub-studies

A total of 284 (142 adolescents and 142 children and their caregivers) participated in the survey (Table 1). We conducted 48 IDIs in total, 8 with children to whom their HIV status had been disclosed, 20 with adolescents and 20 with caregivers of the children (see Appendix 1 & 2 for the characteristics of each). We conducted five FGDs, two with adolescents, two with caregivers and one with children whose HIV status had been disclosed.

### Survey

As shown in Table 1, among children aged 0-14 years included in the survey, 52% were girls with a median age of 9 (IQR 7-12) years. Their main level of education was primary school (87%), and most children were unaware of their HIV status (63%). Among adolescents, 53% were boys, and the median age was 18 (IQR 18-18) years. Their level of education ranged from primary education (21%), secondary education (72%), to a bachelor’s degree (6%). The median age of first diagnosis was 6 (2–10) years, and the median duration of being in HIV care was 12 (IQR 10-15.8) years.

Most participants had personal phones, with 133 (94%) among caregivers and 105 (74%) among adolescents. More adolescents (23%) shared their mobile phones than caregivers (15%). The majority used simple feature phones (72%) and had a good experience with sending (respectively, 78%, 93%), reading (96%, 99%) and receiving SMS (98%, 98%). Thirty percent of caregivers and 27% of adolescents reported having ever experienced some network problems since owning a phone.

We found that 94 (66%) caregivers and 108 (76%) adolescents had people assisting them in reminding them of medication intake time. Among six caregivers and 30 adolescents who often forgot their medication, 5 (83%) caregivers and 28 (93%) adolescents were interested in receiving reminders. They favoured daily reminder SMS before the usual intake time or during the evening.

**Table 1:**
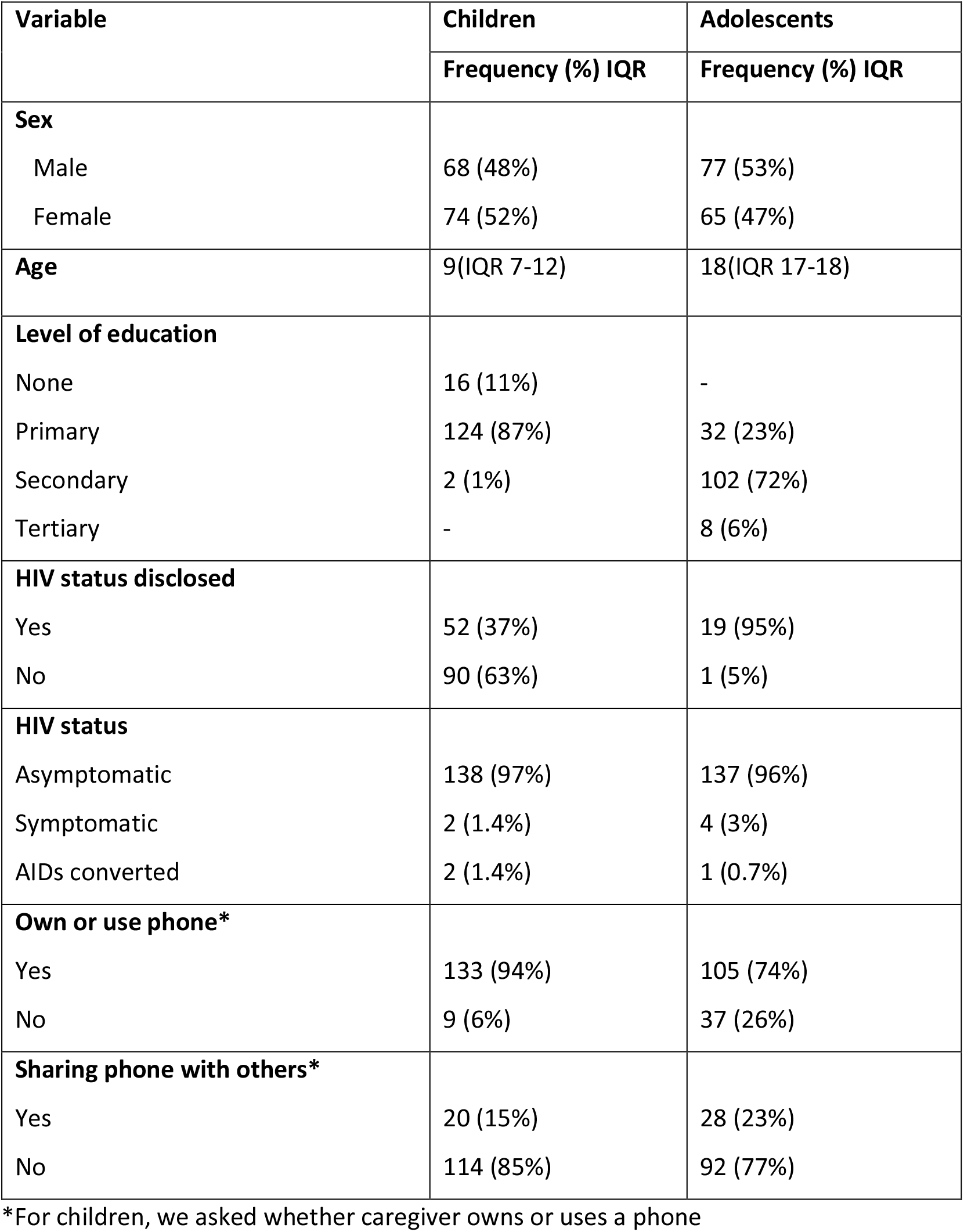
Patient characteristics of children with caregivers (N=142) and adolescents (N=142) participating in the survey.

### Acceptability of DAT

#### Exit interviews, IDI and FGD

We present the results about the preferred SMS contents and the acceptability of the intervention according to the seven constructs of the TFA. We describe SMS content preference under the ‘*affective attitude*’ construct of TFA. Table 2 describes the participants’ general experiences of using the DAT.

**Table 2.** Experience with using DAT as obtained from the exit interviews.

| Variable | Children | Adolescents |
| --- | --- | --- |
|  | Frequency (%) | Frequency (%) |
| **Experience with receiving reminder SMS** |  |  |
| Very good | 10(50%) | 3(15%) |
| Good | 8(40%) | 15(75%) |
| Neutral | 1(5%) | 2(10%) |
| Not good | 1(5%) | 0(0%) |
| **Experienced a mobile Network problem** |  |  |
| Yes | 5(25%) | 4(20%) |
| No | 15(75%) | 16(80%) |
| **Confirms that SMS makes you to take medication** |  |  |
| Yes, always | 16(80%) | 15(75%) |
| Most times | 2(10%) | 5(25%) |
| Mostly not | 2(10%) | 0(0%) |
| **Think of Wisepill box** |  |  |
| It is very nice | 7(35%) | 7(35%) |
| It is nice | 12(60%) | 13(65%) |
| Neutral | 1(5%) | 0(0%) |
| **Concerned about being daily monitored** |  |  |
| Yes | 0(0%) | 1(5%) |
| No | 20(100%) | 19(95%) |
| **Experienced difficulties using the device** |  |  |
| Yes | 1(5%) | 1(5%) |
| No | 19(95%) | 19(95%) |
| **Liked the contents of the feedback** |  |  |
| Yes | 20(100%) | 15(75%) |
| No | 0(0%) | 5(25%) |
| **Understands the graph** |  |  |
| Yes | 18(90%) | 20(100%) |
| No | 1(5%) | 0(0%) |
| **Thinks feedback graph can improve adherence** |  |  |
| Yes | 18(90%) | 19(95%) |
| No | 2(10%) | 1(5%) |

##### Affective attitude

In the exit interviews, 90% of adolescents and children’s caregivers indicated having had a good or very good experience receiving reminder SMSs (Table 2). Participants liked the device, its appearance, shape, and colour, mainly because it was difficult for others to know its function. Hence, it was a helpful tool to safely store medication and preserve confidentiality, especially among participants attending boarding schools, where pupils spend their entire time on campus. Parents/caregivers of adolescents expressed happiness that the device had a monitoring feature to track whether adolescents were taking their medication.

> *“When I was included in this project, I was really happy because I thought it would help me to know when my child has taken his medications and when he hasn’t. So, I know what is causing his disease progress to be bad or good sometimes.” (IDI, caregiver)*
>
> *An adolescent in boarding school explained;*
>
> *“Yes, I liked it much better than the hospital container. I can go with it anywhere; no one notices I carry medication. (IDI, adolescent)*

Moreover, participants shared positive feelings about how the nurses explained the feedback graph (Appendix 5) and gave education on medication intake. All caregivers and 75% of adolescents positively evaluated the graphs’ content. Participants indicated that it provided a clear overview with different colours about medication intake per day. We found similar results in the IDIs in which some participants indicated being worried about living *without* the Wisepill® box. Nevertheless, 25% of adolescents were frustrated with the feedback because it showed that they did not open the box, often due to technical malfunctioning, while they were ingesting pills.

> *“I didn’t like it because there was a time when I was taking medication. However, they [the pills] were kept in another pill bottle than the Pillbox device, so when I came back, I found that my medication intake percentage had decreased.” (IDI, adolescent)*

Most participants liked the SMSs that used general language, did not mention the word ‘medication’ and were not easy for others to understand the purpose of the SMS (Appendix 3 & 4). Among adolescents, the most preferred SMS contents were ‘*Your health is important’* (90%), ‘*The time is at hand’* (95%) and *‘Care for your health’* (95%). Caregivers of children frequently preferred ‘*Do not stop protecting the child’* (80%), *‘Care for the child’s health’* (85%), and ‘*The time for the child to use is at hand’.* (90%).

> *“Okay, ‘remember to consider your health’…. I like it because it gives me the motivation to be taking my medication on time.” (IDI, adolescent)*

Participants disliked receiving the SMS mentioning the word ‘medication’. The specific content of the first SMS, ‘*your time to take medication is near; you are reminded to take your medication as directed by healthcare workers,* was not preferred by 8 (40%) caregivers and 6 (30%) adolescents. The SMS was too long, contained too much information and would raise questions when seen by others.

Data from IDIs and FGDs confirmed these findings. Adolescents had several opinions about SMS. Some suggested that an abbreviation would have been sufficient to make them remember. Some participants were irritated by the content of the SMSs and doubted the overall effect of the reminder SMS; as for them, SMSs just increased the tension of unwanted disclosure of status.

An adolescent in secondary school, said:

> *“Some of the messages were very difficult for me, especially when they came in while I was with friends. If the system of messages could be changed to be ‘Take care’ or ‘Take care of your health ‘or start with ‘Dear Customer…if’. So, when you are with your friend, you could tell him: ‘aah … those are the messages from the network providing company’. This could be better than that of medicine, medicine every time. People could realize that this guy is on medications.” (FGD, adolescent)*

> *“I hate that message [message sent in the first week…], because you find there at times, I have given my phone to a friend, and that message of reminding me to take medication comes in, which says: ‘you are reminded to take your medications’. I shall have to lie to my friend that I had a headache that is why the health experts are reminding me to take medications” (FGD, adolescent)*

Participants also suggested educational topics to be sent in SMS. Adolescents and children that were aware of their HIV status mentioned they would like to receive an SMS on how to prevent HIV transmission to others, information about nutrition, entrepreneurship, reproductive health, and family planning methods. Caregivers wanted education on how they can best disclose to their children their HIV status.

##### Intervention coherence

Participants understood how the device worked. Most participants reported that the device was functioning well, and always received the first reminder SMS in time, and if they opened the device, they did not get the second reminder SMS. Participants found the device easy to open and pack medication. Nevertheless, some participants thought it had cameras so healthcare workers could see them. A child’s caregiver elaborated:

> *“My grandchild would often say: ‘grandma, the message just came in. Where is the device so I take my medication? If I do not take them on time, they will see us.’ I had told her that if she doesn’t take it on time the device takes her picture” (FGD, caregiver)*

A child’s caregiver explained:

> *“When you did not open [the box], you get two SMS. If you open it, the second SMS does not come. I wish that if you gave the child [medication], they would also say, today you have used correctly on time”*.

However, we also found that adolescents often faced difficulties using the Wisepill box appropriately, affecting the results of their adherence levels. A minority of adolescents reported a low battery and having no time to change the device, having lost their mobile phone or forgetting to bring the device during travel. Specifically, in one case, because of a desire to discard the medical pill bottle, an adolescent put all medications in the pillbox, resulting in a situation where the box could not close and communicate properly.

> *“I do open the device, but I take the medication from the can…. because they said, if you open the device, it will count and show the remaining29 pills. Therefore, I take from the can container and sometimes from the device.” (FGD, adolescent)*

Generally, the reminder SMSs were found easy to understand. Adolescents said that since they were told at study entry that they would receive reminder SMS, they were expecting them. When they received the SMS from the study number, they automatically understood that it was for reminder purposes.

> *“The SMSs are easy to read and understand…. I do not use much time. When I read it, I just know it is for reminding medication time” (IDI, adolescent)*

All adolescents and children’s caregivers indicated to have seen and understood the graph during the feedback consultation, and they believed that the nurses explained the graph clearly. They knew how the device communicated about their adherence and how that information appeared in the graph.

> *“Firstly, it is about the use of drugs and the consequences if I do not take them. And then, we go on with other nutritional issues. Second, the nurse often tells me that my adherence is good.” (IDI, adolescent)*.

> *“The graph showed that there was a day that I didn’t take my medication at all and there was a day that I took my medication too late.” (IDI, child)*

##### Perceived effectiveness

Participants considered daily SMS effective. Also, participants mentioned that SMS timing of 30 minutes before medication time was reasonable since it allowed them some preparation time to plan and reach home on time. Nevertheless, others suggested reducing the time to 20 or even 5 minutes closer to intake time to prevent forgetting to take medication, despite receiving the reminder SMS.

The device was considered effective in making participants take their medication on time. This was through reminders which addressed forgetfulness problems, but also the feeling that they were being monitored stimulated them to be adherent to pill intake time. In the exit interviews, none of the participants reported ever having opened the device to satisfy the researchers or to avoid getting the second reminder SMS.

Participants explained how the device assisted them with remembering, for example;

> *“In the past, if you were tired, you would say: ‘I shall take medication tomorrow’, but now you know you are being detected. You just have to take.” (IDI, adolescent)*

> *“My child used to refuse to take the medication, but now when he hears the text messages, he says: ‘that is my message’ and goes to take the device, so I can give him the medication.” (FGD, caregiver)*

Even though the device was perceived effective in reminding medication time, other factors, i.e. a non-confidential environment, the phone being in silent mode, the phone not being charged, or network problems, still played a role in hindering adherence to treatment among participants. Sometimes participants just ignored the SMS, thinking they would take the medications later. Some participants admitted:

> *“Since I got this device, only two days I did not take medicine because the environment I was in at that time was not easy to open it and take medicine, so I waited for that time to pass and took them.” (IDI, adolescent)*

> *“I had my problems and not the matter of forgetting. I was just ignoring it when messages came in. I pretended that I was busy and I didn’t take the medication.” (IDI, adolescent)*

Nearly all adolescents (95%) and 90% of children indicated that the graph assisted them in improving adherence to their medication in the next month. Respondents claimed that the information in the graph increased their motivation and confidence to take medication every day and reach the level of adherence as agreed with the nurse. Interestingly, a few participants mentioned that the device encouraged greater accuracy in discussing their adherence behaviour with the nurse.

> *“I have already seen my medication intake for the past month, so the next month I will try to take my medications better, so those mistakes will not happen again.” (IDI, adolescent)*

##### The burden

Most participants deleted the reminder SMS immediately after reading them, regardless of whether they liked the content or not. The reasons for deleting were to empty phone space for a few participants, but most participants did that to prevent unwanted disclosure.

The fact that the DAT sends reminders through a mobile phone created a challenge for participants without a personal phone. This was more noticeable among participants in boarding schools, as there they are not allowed to have phones at school. In this case, the reminder SMS was received by a school caretaker or a parent/caregiver at home who would then call or forward the SMS to the school’s caregiver. Some wished the device could ring an alarm during medication time instead of sending an SMS through the phone.

A child’s caregiver said:

> *“It would be good if the device could ring an alarm…. that is when the time comes I may be far from home still working and the child is at home. If the device rings an alarm, she could hear it and take medication instead of waiting on me to reach home.”*

Twenty-five percent of caregivers and 20% of adolescents experienced mobile or device network problems (Table 2). Consequently, they would receive the second reminder while they had already opened the device. Some changed their medication intake time without informing the research team, resulting in inadequate SMS traffic.

An adolescent when asked about her experience with the use of DAT responded:

> *“I had a bad experience because you have taken your medication and then they accuse you that you haven’t taken your medication, and the other message comes.”*

A child’s caregiver criticised:

> *“Ever since I received this device, everyday it’s ‘you have not given the child medication, you have not given medication’, while I never skip.” (IDI, caregiver)*

None of the interviewed participants found the feedback consultation too long or that coming to the clinic was a burden to them. We asked participants about being monitored daily, and even though one adolescent (5%) felt anxious (Table 2), most individuals did not share personal concerns about adherence counselling using real-time feedback. Nevertheless, in one of the FGDs, some participants agreed that there were too many questions in a short time.

> *“Anxiety existed within me [about being monitored]. However, I knew it was good because the device was reminding me.” (IDI, adolescent)*

##### Ethicality

Participants considered the device to be user friendly. None of the participants faced stigmatisation because of using the device. None of the participants reported the occurrence of unwanted HIV status disclosure. Some explained that, since the SMS were general and did not mention personal names, even if others happened to see the messages, they could explain that it was not meant for them.

Some of the quotes include:

> *“I have not seen any disadvantage of the device…. Just one! If you do not use it, you will be detected that you did not take your medication.” (IDI, child)*

> *“Only my family saw the SMS, and they find it normal since they know my status. But no one else outside home saw them. I was hiding them from my friends…. friends are not people to trust; they may see it and make you the headline.” (IDI, adolescent)*

##### Opportunity costs

None of the participants incurred additional personal costs, such as costs related to transporting expenditures or time lost time from work to participate in this intervention. The feedback session aligned with their regular appointment visits. Participants only had to return to the clinic in the case of technical device issues and when we asked participants to come to change their devices.

An adolescent explained during an FGD session:

> *“To be very frank ……. the place I am staying, to ask permission every time while I do not want people to know my condition. I don’t like that. So, the device should be adjusted to be stable, so people are not called to the clinic every time it has problems.”*

The intervention worked by sending reminder SMS through a phone. That created a limitation for participants without personal phones. Participants explained:

> *“I see the cost is that you should have a phone since the device doesn’t ring an alarm to say it will remind you.” (FGD, caregiver)*

> *“I didn’t incur any additional costs. Also, when I come to the clinic for my regular appointments, I spend 2000 shilling (about €0,80). It is just 1000 to come here and 1000 back. It doesn’t take too long from where I live.” (IDI, adolescent)*.

## Discussion

This formative study aimed to assess the needs, contents, and acceptability of the DAT with reminder cues and tailored feedback on adherence among CALHIV. We found that CALHIV need DATs and they find it highly acceptable. Most participants had a positive experience with using the device and preferred neutral SMS content that did not mention the word ‘medication.’ Feedback on adherence inspired good adherence behaviour.

### Need for reminders

Results from the survey indicate that children’s caregivers and adolescents who had no people to remind them about the medication time, needed reminders. They were interested in receiving daily reminder SMS before the usual medication time or during the evening. Participants in the DAT intervention confirmed this. Similar results were found in a study done in Peru, where participants preferred text messages over recorded voice messages or phone calls (38). Participants found text messages easier, readily accessible, and more confidential, although they suggested to change the SMS content daily or on a weekly basis to avoid boredom. In another study in Uganda, scheduled daily messages were preferred as they reduced the chance of missing doses compared to weekly or triggered SMS.(47) However, some participants in that qualitative study were worried that frequent text messages for each dose would lead to phone dependency for medication intake, causing more problems in case of phone malfunction.(48)Furthermore, a meta-analysis of randomized control trials reported that scheduled daily text message reminders improved adherence outcomes.(49)

### SMS contents

Preference of SMS content depended on whether participants had disclosed their HIV status to people around them when receiving the SMS and whether they shared their phones. This was also reported by studies conducted in Kenya and Uganda, in which open contents SMS were found frustrating and increased the risk of unwanted disclosure of HIV status.(47,48) Some suggested using code words or setting passwords, but the latter depends on the type of phone owned.(38, 52) The ability of the SMS to preserve confidentiality and privacy was the most critical factor discussed by participants for the acceptability of the SMS based mobile health intervention.

Similar to our previous acceptability studies among PLHIV, participants liked the SMS contents that were motivating in nature, because they made them feel seen and cared for.(30,47,48) Also, participants suggested several topics they would like to receive educational messages about, for example receiving education on how to disclose HIV status to a child. This indicates that participants consider mobile health interventions promising and suitable for addressing HIV-related issues if they preserve confidentiality.

### Acceptability of the intervention

This study found that an SMS-based digital adherence tool with personalized adherence feedback was needed and highly acceptable among CALHIV. Considering *affective attitude*, we found that most participants were satisfied with the intervention. In other participants also described that the device was easy or convenient to use and convenient in preserving confidentiality since it didn’t resemble the traditional pill container.(27, 47, 51) Additionally, in our study, we found that this was particularly important among boarding school adolescents. A study in Kenya among boarding school students reported fear of unintended disclosure due to drug packaging as one of the barriers of ART adherence.(15) Therefore, electronic monitoring devices have the potential to solve these adherence challenges.

DAT was *perceived as effective* in reminding medication time and making participants take their medication on time. It was also explained by participants in other studies that DAT trained them in developing good adherence behaviour in taking the pills daily and on time.(27, 55, 56)

Few adolescents were concerned about being daily monitored. However, their caregivers liked the monitoring feature of the tool as they were able to know if their children took medication or not. A qualitative study, exploring the perceptions on the effect of electronic monitoring tools on adherence, explained that even though electronic monitors created a sense of pressure by forcing participants to adhere, most participants found it beneficial in assisting them in reaching their adherence goals and live a healthy life.(54)

Furthermore, we found that most participants understood how the DAT works and how it could improve adherence (*intervention coherence*). . In other, similar studies exploring the acceptability of DAT devices, participants explained good understanding on how to open the device, fill it with medication and record missing doses if the device was not opened.(47, 54) Nevertheless, a few participants described not using the intervention as intended. They packed more medication that could fit in the device, and some swallowed medication from the packaging container instead of from the Wisepill box. Future studies should make efforts to properly train users how the intervention works and insist to only take pills from the device.

All participants received and understood the feedback on adherence graphs as discussed with nurse counsellors. Feedback on adherence motivated them to adhere better to medication. Participants knew that objective adherence information can improve accurate HIV counselling, where nurses can no longer be misled. Objective monitoring has the potential to overcome social desirability bias, which is a well-recognized bias during patient recall. (56)

However, still 25% of adolescents felt dissatisfied about the feedback. This was often due to inaccurate use of the pillbox during the intervention. Some adolescents received lower than expected percentages, but they recalled having taken all pills. These contradicting instances may hamper patient-clinician trust that could be perceived as “not being believed”. Therefore, caution should be exercised when choosing real-time medication monitor (RTMM) adherence results as the new gold standard, but rather frame RTMM as a complement to patient self-report.(57)

Regarding factors affecting acceptability negatively, we found that the intervention could cause *burden* to participants in case of malfunctioning due to technical or network issues. No signal is sent when there is no network, meaning the system believes no medication was taken, so the second reminder was sent. That created a burden to participants when they kept receiving the second reminder SMS and considered to be accused of not taking their medication while in fact they had. For longer period interventions, closer and regular follow-up of participants is needed to detect devices that do not communicate and replace them.

We observed other *burden* issues when participants shared phones or did not have access to phones, like students in boarding schools. Future interventions should include reminders features such as device alarms to assist participants with phone challenges.

Nevertheless, SMS-based interventions requires participants to have phones, be literate and have access to electricity and a good network connection.(35, 39) Consequently, these interventions are not always accessible to the people who need it. Other reported barriers include battery life, power failures and potential stigmatization due to DAT.(44,51,58) These studies similarly reported that fear of unwanted disclosure of HIV status led participants to delete the SMS shortly after reading them, and some could not take their medications even after reminders due to lack of privacy or psychosocial factors.

Participants suggested to receive educational SMS on different topics, such as prevention of HIV and how best to disclose the HIV status to a child or a partner. However, this should consider the risk of unwanted disclosure, hence careful phrasing of HIV-related health education is important and should be tailored based on recipient’s desires.(36, 39)

## Study Limitations and Strengths

Due to the nature of the intervention, only literate participants could participate. Future DAT studies should consider interventions suitable to participants that cannot read, for example, using devices with alarms. A second limitation of this study was the small sample size of participants involved in the DAT study and the duration of one month of using the intervention. Therefore, caution should be taken when generalizing study results.

The study strengths include that it focused on vulnerable key populations, i.e. CALHIV while most studies on DAT have been among adults. This study provided an opportunity for CALHIV to give their opinion and shape a DAT intervention suitable to them and to address their unique challenges.

Also, we used purposive sampling to select participants for the DAT intervention. Attempts were made to approach a diverse set of adolescents. However, it was hard to establish a representative heterogeneity for age. More than half of our participants were 18 years or older. This skewed distribution can be attributed to the fact that informed consent was legally required from parents of underage adolescents. In Tanzania, it is common for young adolescents to visit the clinic alone without their parents, hindering getting consent from their parents. As such, it was difficult to recruit participants below 18. We made efforts to reach the parents by phone and asked them to visit the clinics to receive more information about the study and, if possible, provide consent for their children to participate.

## Conclusion

DAT with reminder SMS and tailored feedback was acceptable among CALHIV. Participants preferred more neutral wording in SMS. We will use preferred SMS in a future clinical trial, which will assess the effectiveness of DAT in improving adherence among CALHIV. Future studies should consider measures to solve network problems such as increasing the period between the first and second SMS, improvements on the device to include alarms for participants lacking phones, and improvements on training about how to properly use the device.

## Data Availability

The quantitative and qualitative data used in this study will be made available upon request from the corresponding author.

## Acknowledgements

We would like to thank all children and adolescents, together with their caregivers for taking part in this informative study. Their opinions and ideas shape the future of our interventions to benefit the general society. We thank the health care workers in participating facilities and the KCRI REMIND team staff who helped to collect data. Lastly, we thank the funder, the European and Developing Countries Clinical Trials Partnership (EDCTP) for supporting our work under the senior fellowship plus TMA2818.

**Appendix 1:** Characteristics of children in the DAT intervention (N=20)

| Sex | Age range | Years in HIV care | Self-reported Adherence (%) | Pharmacy refill Adherence (%) | DAT Adherence (%) |
| --- | --- | --- | --- | --- | --- |
| Male | 6-10 | 7 | 100 | 100 | 96 |
| Male | 11-15 | 9 | 100 | 79 | 100 |
| Male | 11-15 | 10 | 100 | 100 | 85 |
| Male | 6-10 | 7 | 100 | 82 | 97 |
| Female | 6-10 | 4 | 100 | 100 | 100 |
| Male | 11-15 | 0 | 100 | 97 | 100 |
| Male | 0-5 | 1 | 100 | 100 | 100 |
| Female | 11-15 | 9 | 100 | 100 | 100 |
| Male | 11-15 | 9 | 100 | 100 | 100 |
| Female | 6-10 | 7 | 100 | 42 | 100 |
| Female | 6-10 | 2 | 100 | 42 | 96 |
| Female | 11-15 | 3 | 100 | 100 | 100 |
| Male | 11-15 | 9 | 100 | 10 | 96 |
| Female | 6-10 | 5 | 100 | 80 | 80 |
| Male | 11-15 | 9 | 100 | 100 | 97 |
| Male | 6-10 | Unknown | 100 | 100 | 08 |
| Female | 6-10 | 10 | 100 | 91 | 32 |
| Female | 6-10 | 3 | 100 | 86 | 100 |
| Female | 0-5 | 0 | 100 | 100 | 100 |
| Male | 11-15 | 9 | 67 | 96 | 73 |
| Median (IQR) | 10(7.2-11.8) | 7(2.2-9) | 100(100-100) | 98.4(80.5-100) | 98.5(87.7-100) |

**Appendix 2:** Characteristics of adolescents in the DAT intervention (N=20)

| Sex | Age range | Years in HIV care | Self-reported Adherence (%) | Pharmacy refill Adherence (%) | DAT Adherence (%) |
| --- | --- | --- | --- | --- | --- |
| Male | 15-19 | 6 | 93 | 98 | 0 |
| Female | 15-19 | 17 | 97 | 82 | 20 |
| Female | 15-19 | 12 | 100 | 92 | 56 |
| Female | 15-19 | 17 | 100 | 53 | 47 |
| Male | 15-19 | 16 | 90 | 87 | 24 |
| Male | 15-19 | 11 | 90 | 90 | 20 |
| Female | 15-19 | 10 | 100 | 100 | 92 |
| Male | 15-19 | 15 | 93 | 79 | 4 |
| Male | 15-19 | 06 | 100 | 97 | 100 |
| Female | 15-19 | 16 | 100 | 99 | 79 |
| Female | 15-19 | 12 | 100 | 100 | 50 |
| Male | 15-19 | 10 | 100 | 97 | 100 |
| Male | 15-19 | 08 | 100 | 100 | 84 |
| Male | 15-19 | 16 | 87 | 100 | 96 |
| Male | 15-19 | 12 | 100 | 100 | 67 |
| Female | 15-19 | 12 | 100 | 100 | 25 |
| Male | 15-19 | 13 | 77 | 100 | 76 |
| Male | 15-19 | 13 | 100 | 97 | 88 |
| Male | 15-19 | 10 | 100 | 90 | 89 |
| Female | 15-19 | 12 | 100 | 94 | 100 |
| Median (IQR) | 18(18-18) | 12(10-15.8) | 100(93.3-100) | 96.9(90-100) | 72(24.2-91.2) |

**Appendix 3:** Adolescents SMS Preference (N=20)

|  | SMS contents | YES (%) | No (%) | Do not remember the SMS (%) |
| --- | --- | --- | --- | --- |
| WEEK1 | "Hello, your time to take the medication is near, you are reminded to take your medication on time as directed by the health care workers" | **13(65%)** | 6(30%) | 1(5%) |
| WEEK2 | "Remember to observe your health."<br>"Remember to protect your health today."<br>"Don't stop caring for your health today"<br>"Your time is at hand"?<br>"You are reminded to protect your health"<br>"Remember to drink on time"?<br>"Your time to use is at hand" | 16(80%)<br>16(80%)<br>18(90%)<br>15(75%)<br>17(85%)<br>15(75%)<br>12(60%) | 3(15%)<br>3(15%)<br>1(5%)<br>4(20%)<br>2(10%)<br>4(20%)<br>6(30%) | 1(5%)<br>1(5%)<br>1(5%)<br>1(5%)<br>1(5%)<br>1(5%)<br>2(10%) |
| WEEK3 | "You are reminded to drink"?<br>"Do not forget to use"?<br>"The time is at hand"?<br>"Your health is important"<br>"Do not forget to protect yourself"<br>"Care for your health"<br>"Drinking is caring"! | 14(70%)<br>16(80%)<br>19(95%)<br>18(90%)<br>18(90%)<br>19(95%)<br>9(45%) | 4(20%)<br>2(10%)<br>0(0%)<br>0(0%)<br>1(5%)<br>1(5%)<br>4(20%) | 2(10%)<br>2(10%)<br>1(5%)<br>2(10%)<br>1(5%)<br>0(0%)<br>7(35%) |
| WEEK4 | "Remember"?<br>"Use"?<br>"Take care"?<br>"Drink"?<br>"Health"<br>"Care for your health"<br>"Value health"? | 13(65%)<br>13(65%)<br>15(75%)<br>15(75%)<br>15(75%)<br>18(90%)<br>15(75%) | 1(5%)<br>2(10%)<br>1(5%)<br>4(20%)<br>1(5%)<br>1(5%)<br>1(5%) | 6(30%)<br>5(25%)<br>4(20%)<br>1(5%)<br>4(20%)<br>1(5%)<br>4(20%) |

**Appendix 4:**
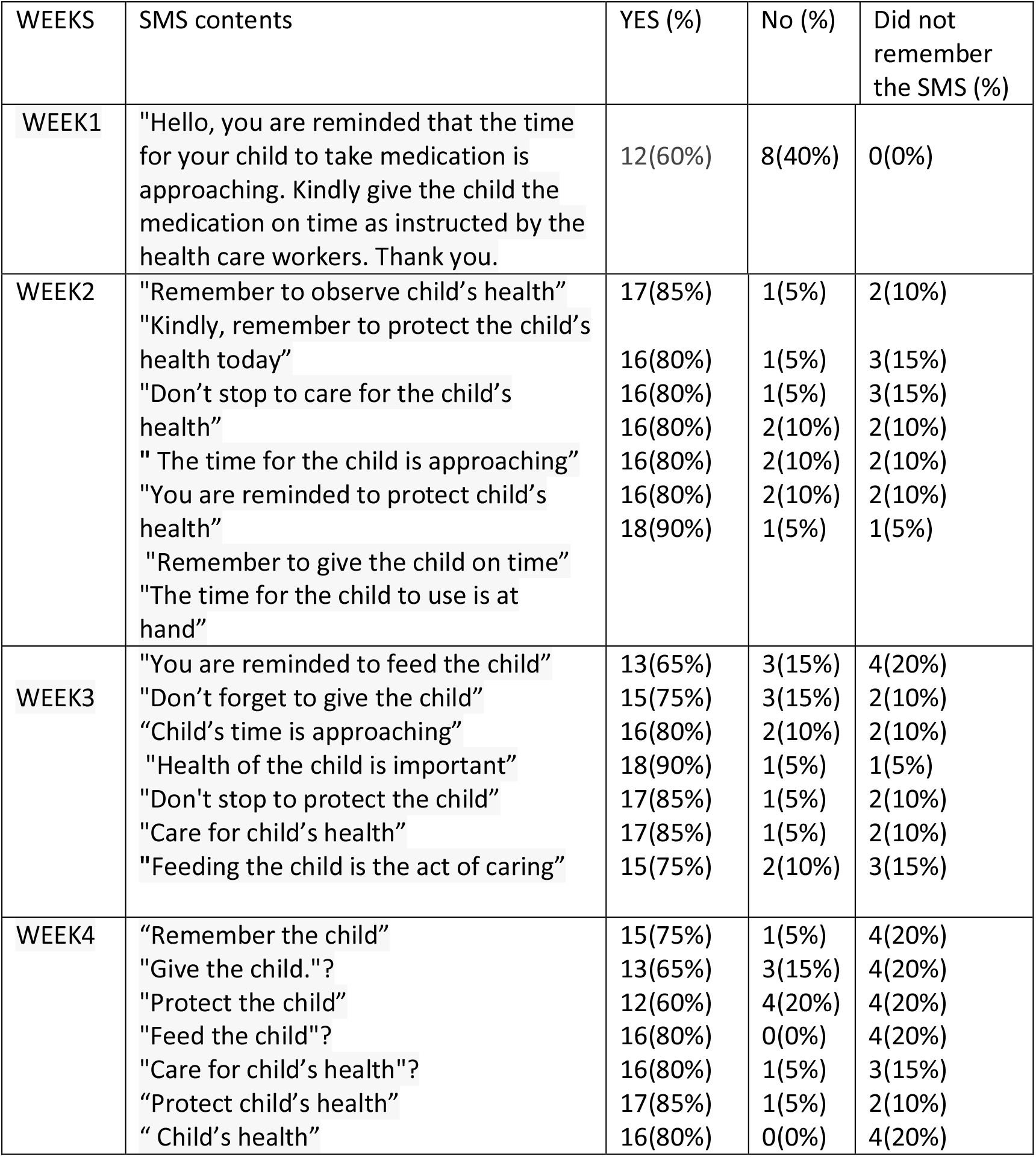
Children SMS Preference (N=20) (Questions were answered by caregivers/parent)

**Appendix 5:**
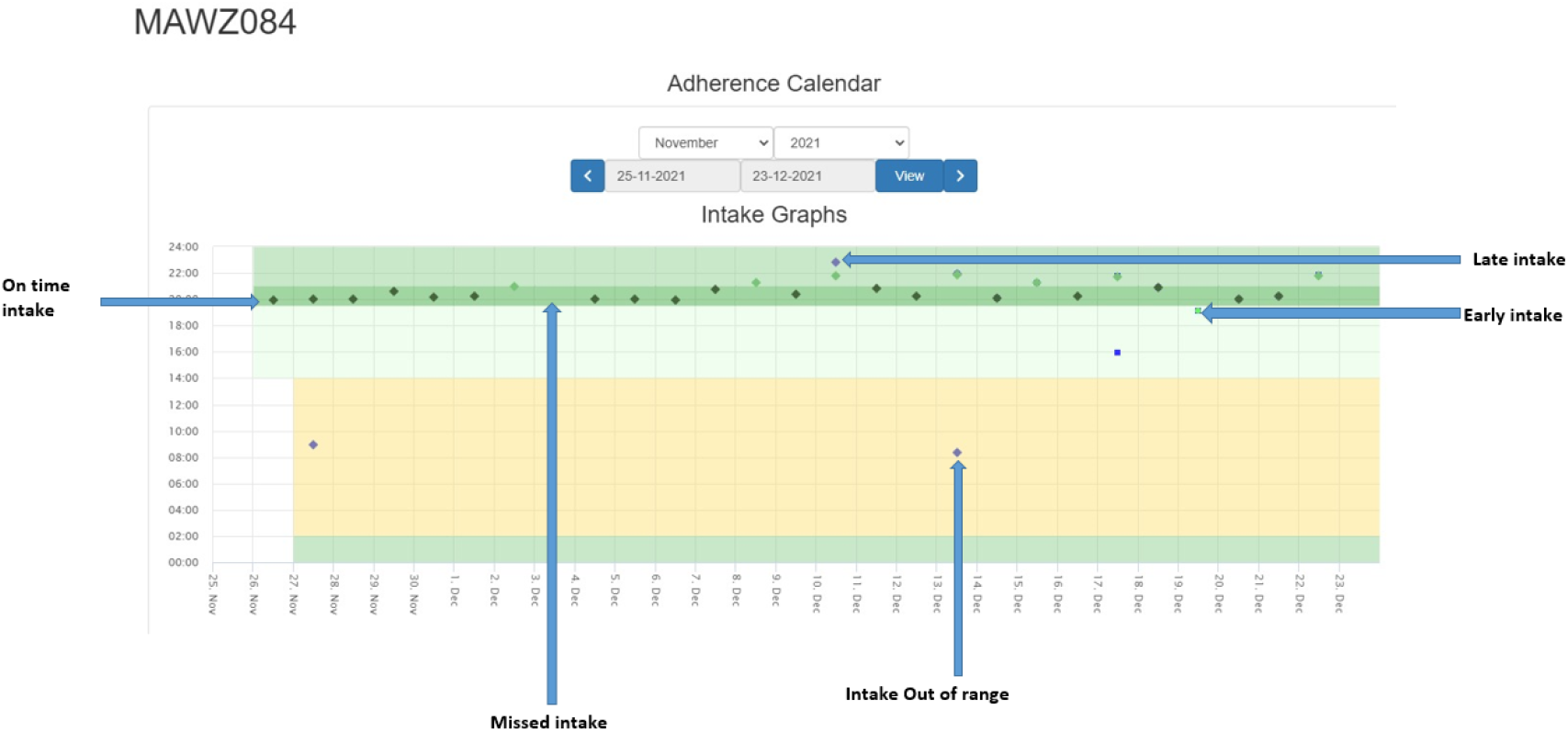
Example of an adherence feedback graph.

## Notes

### Competing Interest Statement

The authors have declared no competing interest.

### Funding Statement

This study was funded and supported by the European and Developing Countries Clinical Trials Partnership (EDCTP) under the senior fellowship plus TMA2818.

### Author Declarations

The study was approved by the College Research and Ethical Review Committee (CRERC) of Kilimanjaro Christian Medical University College (KCMUCo) and the National Health Research Ethics Sub-Committee (NatHREC) of the National Medical Research Institute (NIMR) of Tanzania.

